# Assessing and predicting neuropathic pain after spinal cord injury: a TRACK-SCI study

**DOI:** 10.1101/2021.11.30.21267098

**Authors:** Kenneth A. Fond, Abel Torres-Espin, Austin Chou, Xuan Duong Fernandez, Sara L. Moncivais, J. Russell Huie, Debra D. Hemmerle, Anastasia V. Keller, Vineeta Singh, Lisa U. Pascual, Anthony M. DiGiorgio, John F. Burke, Jason F. Talbott, William D. Whetstone, Jonathan Z. Pan, Phil R. Weinstein, Sanjay S. Dhall, Adam R. Ferguson, Jacqueline C. Bresnahan, Michael S. Beattie, Nikos Kyritsis

## Abstract

Neuropathic pain is one of the most common secondary complications occurring after spinal cord injury (SCI), and often surpasses motor and sensory deficits in the patient population preferences of the most important aspects to be treated. Despite the better understanding of the molecular and physiological mechanisms of neuropathic pain, reliable treatments are still lacking and exhibit wide variations in efficiency. Previous reports have suggested that the most effective pain management is early treatment. To this end, we utilized the TRACK-SCI prospective clinical research database to assess the neuropathic pain status of all enrolled patients and identify acute care variables that can predict the development of neuropathic pain 6- and 12-months post SCI. 36 out of 61 patients of our study cohort reported neuropathic pain at the chronic stages post SCI. Using multidimensional analytics and logistic regression we discovered that (1) the number of total injuries the patient sustained, (2) the injury severity score (ISS), (3) the lower limb total motor score, and (4) the sensory pin prick total score together predict the development of chronic neuropathic pain after SCI. The balanced accuracy of the corresponding logistic regression model is 74.3%, and repeated 5-fold cross validation showed an AUC of 0.708. Our study suggests a crucial role of polytrauma in chronic pain development after SCI and offers a predictive model using variables routinely collected at every hospital setting.

## Introduction

Spinal Cord Injury (SCI) is a catastrophic and untreatable condition and although it occurs in a small percent of the US population, the effects can be devastating. As of 2020, there are approximately 288,000 individuals with SCI currently living in the US with an estimated 17,700 new cases of SCI in the US each year.^1-3^ SCIs include the primary injury, which causes extensive neuronal cell death with immediate consequences, as well as secondary complications or ‘secondary injuries’ that result from inflammation, excitotoxicity, and loss of sensory input below the injury level. These secondary injuries typically arise in the later sub-acute to chronic phases. Due to the relatively small amount of SCI secondary injury studies, organizations such as The North American Spinal Cord Injury Consortium (NASCIC) urge researchers studying SCI to “focus their research efforts on identifying avenues to reduce the impact” of pain among other secondary injury characteristics.^4, 5^

Neuropathic pain, a subcategory of chronic pain, is one such secondary injury known to persist well into the chronic phase and can thus negatively impact a patient’s quality of life. Chronic pain after SCI can be broadly subcategorized into two categories: nociceptive pain that arises from firing of pain receptors, and neuropathic pain as a result of damage to the nerves of the somatosensory system. Previous studies predicted a median prevalence rate of 53% for SCI patients within 1 year of injury with more recent studies showing similar prevalence rates.^6-9^ A 2012 study reported 17% of surveyed patients with neuropathic pain rated their quality of life as being “worse than death”.^10^ Other surveys of SCI patients indicate managing and/or dealing with chronic pain in general ranks third after regaining sexual function or function of upper extremities but ranks first when specified to challenges faced on a daily-basis.^11^ Due to the low prevalence of SCI, most of what is known about neuropathic pain comes from diabetes mellitus patients diagnosed with central neuropathic pain.^12, 13^ Even so, its pathophysiology remains poorly understood and can be difficult for clinicians to identify, much less treat.^6, 13-15^ Common descriptors for neuropathic pain: “burning, uncomfortable cold, prickling, tingling, pins-and-needles, stabbing, shooting, lancinating, tight, swollen, and squeezing sensations that are distressing.”

Treatment for neuropathic pain follows the following criteria: (1) remove underlying cause, (2) provide relief of neuropathic pain symptoms, and (3) prevent neuropathic pain symptoms from interfering with patient’s quality of life. Current treatment schemes include physical therapy, anti-seizure or antidepressant medications, opioids, nerve blocks, peripheral nerve stimulation, brain stimulation, or more invasive procedures.^16^ There is also multimodal therapy where two or more treatments are used in combination to treat and alleviate neuropathic pain symptoms.

Unfortunately, these treatment options for chronic neuropathic pain vary in effectiveness and side-effects between patients.^16-19^ Previous studies have shown early diagnosis and treatment have the potential to reduce the symptoms of chronic neuropathic pain and mitigate the risk of progressing to later stages where advanced treatment with more significant side-effects is required.^20^ To facilitate early diagnosis or treatment, biomarkers able to predict the development of neuropathic pain play a key role.

Most clinical literature with the keywords “neuropathic pain”, “SCI”, “predictors”, and “biomarkers”, follow an approach where researchers primarily select variables to be tested as biomarkers based on domain expertise.^2, 12, 17-19^ To our knowledge, not as many clinical studies follow an unsupervised approach, such as the one used in our study.^13^ Here, the variables analyzed in order to create a predictive model for the development of neuropathic pain come from the TRACK-SCI database including up to 2,000 records per patient during their hospital stay, to develop a predictive model for the development of neuropathic pain.^21^ Critically, initial analysis and selection of the predictive variables was performed in an unsupervised data-driven fashion with guidance from clinical domain expertise as needed. The ability to predict chronic neuropathic pain will help clinicians to initiate preemptive treatment regimens earlier and educate their patients and/or refer them to outpatient specialized services.

## Results

Transforming Research and Clinical Knowledge in Spinal Cord Injury (TRACK-SCI) is an IRB approved multi-center prospective SCI study.^21^ Our first goal was to utilize the large TRACK-SCI database, which includes 135 enrolled SCI patients and up to 22,000 data points for each patient, to assess the chronic pain status of these patients and compare our findings with other similar studies. All the pain data collected in TRACK-SCI are self-reported. Specifically, the questionnaire we administer to the enrolled patients are (1) the International Spinal Cord Injury Pain Basic Data Set (ISCIPBDS) Version 2.0 and (2) the Douleur Neuropathique en 4 Questions (DN4) as our Pain Questionnaire Dataset.^22, 23^ These CRFs were administered over the phone at 3-, 6-, and 12-months post SCI. At the time of this analysis, TRACK-SCI had 136 enrolled patients. For 3 months post SCI, 4 patients withdrew or were deceased, 1 was not yet due for a follow-up, and 73 did not have a completed follow-up. For 6 months post SCI, 8 patients withdrew or were deceased, 2 were not yet due for a follow-up, and 72 did not have a completed follow-up. For 12 months post SCI, 10 patients withdrew or were deceased, 6 were yet due for a follow-up, and 73 did not have a completed follow-up. In the end, 58, 54, and 47 patients remained at 3, 6, and 12-months respectively (**Fig. 1**). The initial summarization of the data revealed that patient retention is a significant challenge for SCI prospective clinical studies at the chronic phase and is especially true for settings like Zuckerberg San Francisco General Hospital and Trauma Center (ZSFG) where rehabilitation always continues at different facilities.

**Figure 1.**
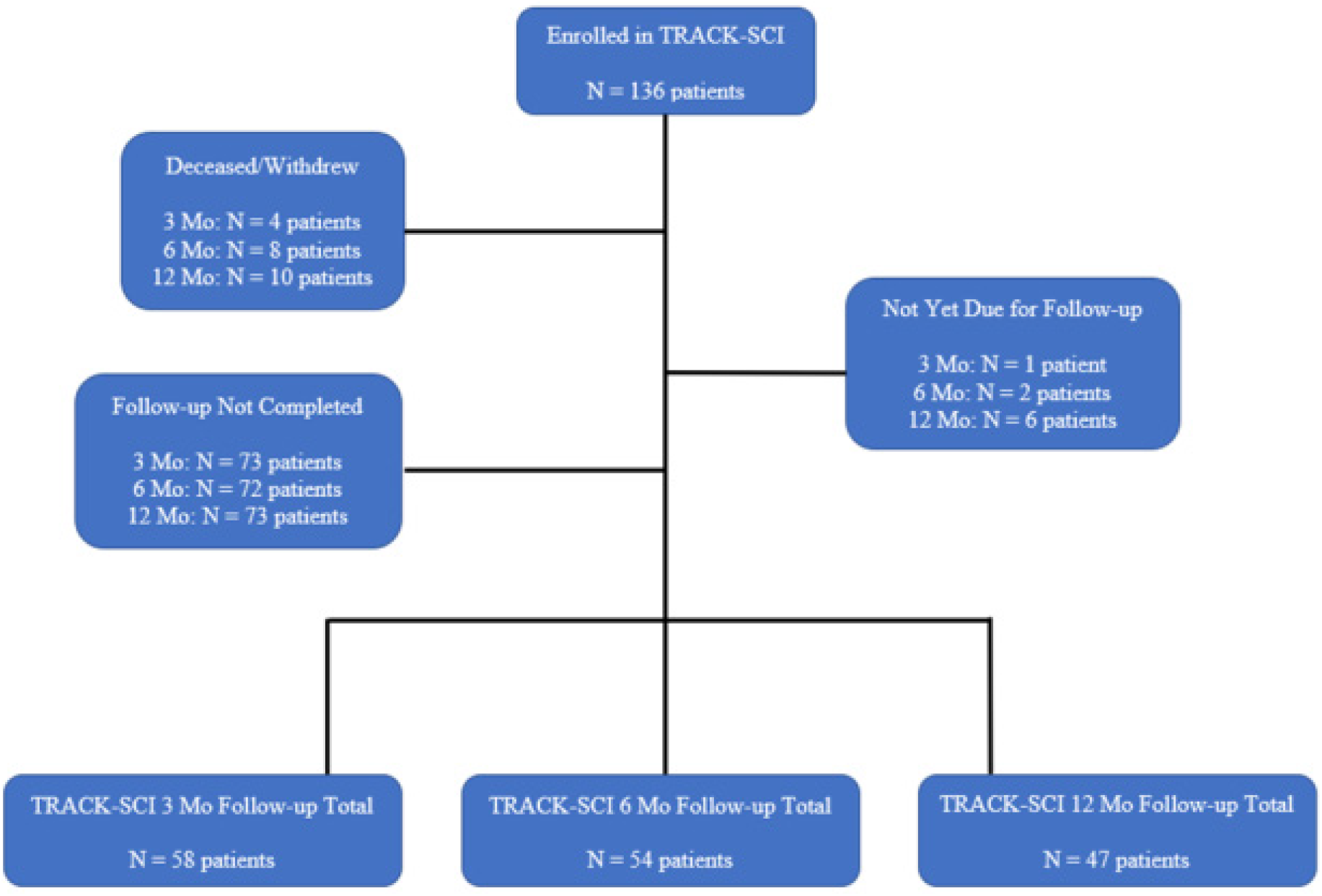
TRACK-SCI enrolled patients flow chart. By the time this analysis started, 136 SCI patients had enrolled the TRACK-SCI study. Patients were followed chronically after SCI, at 3-, 6-, and 12-months post injury and their pain status was determined through self-reported questionnaires. As it often happens in clinical studies the drop-out rate was significant with 58, 54, and 47 completed pain questionnaires at 3-, 6-, and 12-months post injury respectively.

Next, we examined deeper the pain status of our enrolled patients using the pain CRFs separately at each time point. First, we observed that for all three time-points pain was reported from over 80% of the SCI patients (89.7%, 100%, and 80.9% for 3-, 6-, and 12-months post SCI respectively; **Fig 2A**). Subsequently, and knowing the importance of neuropathic pain in the overall quality of life of SCI patients, we used the CRF data to categorize the pain into ‘nociceptive’, ‘neuropathic’, and ‘both’. We could confirm what was shown by previous studies before^16, 24^ that neuropathic pain is the main pain problem the SCI patients suffer from as it was present in 75%, 63%, and 71.1% of our SCI cohort at 3-, 6-and 12-months post SCI respectively (**Fig. 2B**). Moreover, we assessed how many different neuropathic pain areas our enrolled SCI patients reported. The vast majority of them reported only one neuropathic pain area (74.4%, 70.6%, and 70.4% at 3-, 6-, and 12-months post SCI respectively) with a few of them reporting up to three areas of neuropathic pain (**Fig. 2C**). For each one of the neuropathic pain areas, the SCI patients were asked to describe the intensity with scores from 0 (no pain) to 10 (pain as bad as you can imagine). There is a statistically significant difference across the three timepoints (5.46 ± 2.18, 4.19 ± 2.91, and 5.93 ± 2.94 for 3-, 6-, and 12-months post SCI respectively), mainly driven by the 6-month time point where slightly reduced neuropathic pain intensity was reported (One-way ANOVA p-value = 0.02). However, the fact that the overall distributions appear similar, and the number of cases vary across the timepoints, do not allow more in-depth interpretations of these data (**Fig. 2D**). Together, these data show the pain manifestation in the TRACK-SCI enrolled patients over time suggesting minor differences in the type and intensity.

**Figure 2.**
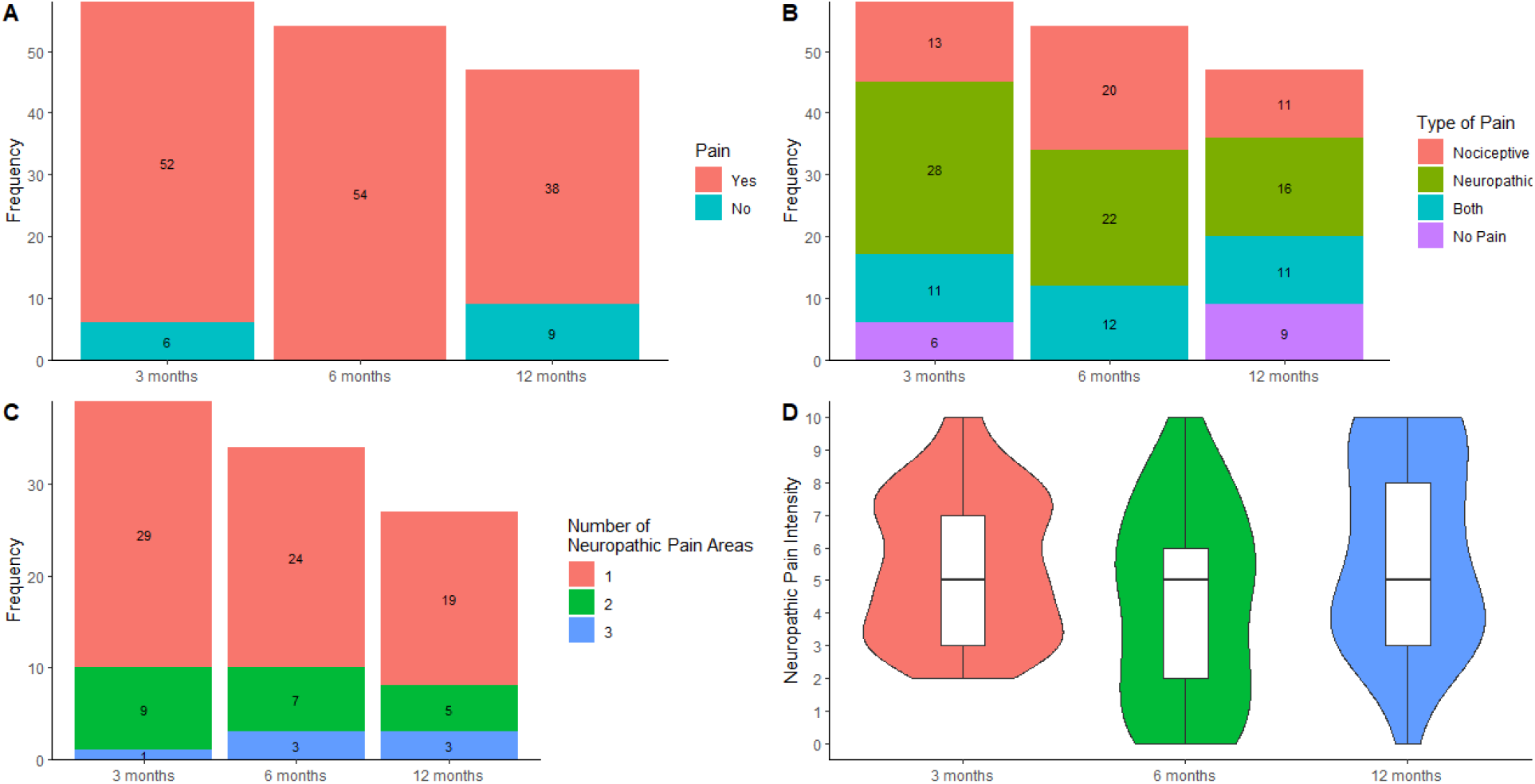
Chronic pain assessment of TRACK-SCI enrolled patients. A. The vast majority of SCI patients experience pain at 3-, 6-, 12-months post SCI. **B**. From the SCI patients who reported pain in A, the most prevalent type of pain is neuropathic across all time points with a significant share of them reporting both neuropathic and nociceptive pain. **C**. Most patients experiencing neuropathic pain post SCI reported that the pain was confined in one area, with few of them listing two areas and a few cases with three neuropathic pain areas. **D**. The average pain intensity of the neuropathic pain areas does not change significantly over time.

One of the major goals of our study is to determine whether we can predict the development of neuropathic pain after SCI using data collected acutely and during their initial stay at the hospital. Because neuropathic pain usually appears within the first 6 months post SCI, we decided to use only the data from the 6- and 12-months post SCI questionnaires. To increase our sample size, we collapsed the 6- and 12-month data into one while using the latest data point for the cases where we had data for both. Collapsing these two time points resulted in a cohort of 61 patients with pain data. We extracted the acute care data we had for these patients from the TRACK-SCI database. The initial dataset included 2,047 variables. Through data-driven methods and domain expertise (see Materials and Methods) we narrowed our dataset number of variables to 63 (**SFig. 1**). Even though the missingness of the data was addressed significantly during our variable selection scheme, there were still missing data in our final dataset. While a small percentage of missing data could be allowed for some analyses, we required a complete dataset and thus imputed the missing data points using multiple imputation by chained equation.

We then used principal component analysis (PCA), an unsupervised multivariate technique, to determine whether any principal component or a combination of them could be used as predictors for chronic neuropathic pain development after SCI. First, we performed a nonlinear PCA and a permutation test (1,000 permutations) to determine how many principal components (PCs) to use in downstream analyses. The permutation results showed that the first 5 PCs are stable and significantly different from random noise (**Fig. 3A**). These PCs together account for 54.4% of the total variance. Using another permutation test for each individual variable, we determined which variables were significantly contributing (i.e. loading) to each one of these PCs (**SFig. 2**). The first two PCs (28.9% of total variance), the ones with the most variables significantly contributing are visualized **Fig. 3B-C**. PC1 represents an overall clinical picture of the patients in the emergency department, while PC2 appears to reflect on pain severity and sensation.

**Figure 3.**
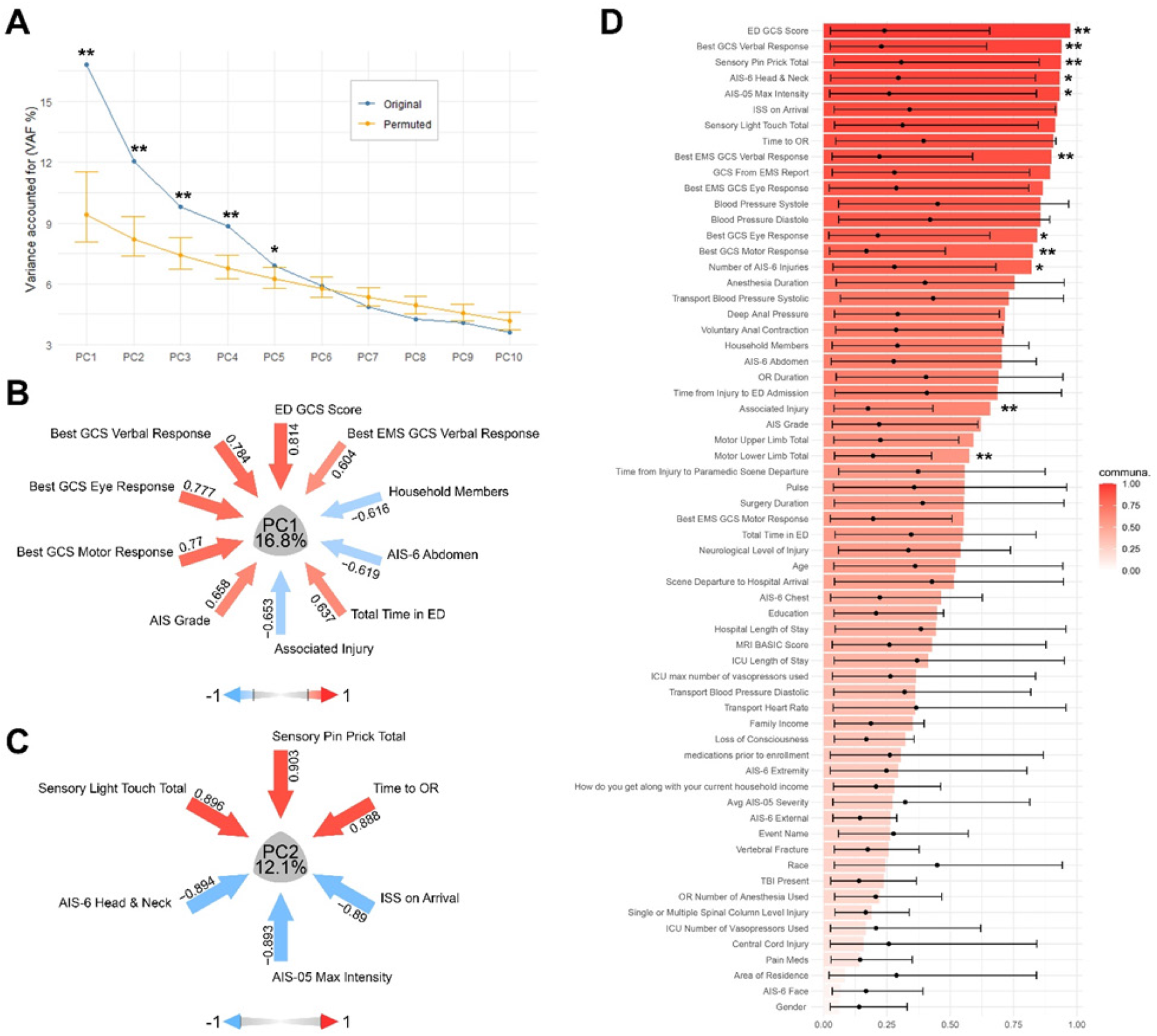
Principal Component Analysis and permutation analysis is used to rank the acute care variables. **A**. Nonlinear PCA analysis on the 63 acute care variables followed by 1,000 permutation tests reveal 5 stable Principal Components that account for 54.4% of the total variance. **B-C**. The highest loading variables in PC1 portraits the ED clinical picture of the patients and while the PC2 variables represent pain severity and sensation. **D**. One thousand permutation tests on the communalities of the first 5 PCs is used to rank the importance of the variables and measure their stability. * p < 0.05, ** p < 0.01

However, using these two PCs alone, together, or in various combinations with the remaining three PCs as predictors in a PC-only logistic regression model was not sufficient to accurately predict the development of neuropathic pain (data not shown). Since the PCs were not good predictors, we decided to use the PCA outcome as a method to perform unsupervised variable selection. We calculated the communalities of each variable in the first 5 PCs (**Fig. 3D**) and assessed their significance above random chance by permutation test of 1,000 permutations. We then selected the important variables (p < 0.1, P = 20) from the permutation of communalities to be used in a logistic regression model to predict the presence of neuropathic pain.

As mentioned, we have pain data at chronic time points (6- and/or 12-months post SCI) from 61 patients, 36 of which reported neuropathic pain (**Fig. 4A, Table 1**). Stepwise forward and backward logistic regression was used to determine the optimal model. The final model includes four variables: (1) the total number of AIS-6 injuries diagnosed in the ED, (2) the Injury Severity Score (ISS) at hospital arrival, and (3) the Sensory (Pin Prick) and (4) Motor (Lower Limb) Total Scores from the ISNCSCI examination performed at the day of hospital discharge (**Fig. 4B-E**). The overall accuracy of the model is 75.4% (balanced accuracy 74.3%) with a p-value of 0.006 (**SFig. 3**). A caveat of our model is that it has not been tested in an independent, external dataset which makes it difficult to assess its validity and the possibility of overfitting to the data. To mitigate this, we performed 5-fold cross-validation, repeated 1,000 times. While our full model had an AUC of 0.793, the repeated cross-validation gave an average AUC of 0.708 (**Fig. 4F**).

**Table 1.** Demographic and important clinical information of the 61 patients with complete pain data at 6- and/or 12-months post SCI.

|  | Yes<br>(N=36) | No<br>(N=25) | Total<br>(N=61) |
| --- | --- | --- | --- |
| **Sex** |  |  |  |
| Female | 11.0 (30.6%) | 4.00 (16.0%) | 15.0 (24.6%) |
| Male | 25.0 (69.4%) | 21.0 (84.0%) | 46.0 (75.4%) |
| **Race** |  |  |  |
| American Indian or Alaska Native | 1.00 (2.8%) | 0 (0%) | 1.00 (1.6%) |
| Asian | 7.00 (19.4%) | 7.00 (28.0%) | 14.0 (23.0%) |
| Black or African-American | 6.00 (16.7%) | 2.00 (8.0%) | 8.00 (13.1%) |
| Hispanic | 4.00 (11.1%) | 3.00 (12.0%) | 7.00 (11.5%) |
| Native Hawaiian or Other Pacific Islander | 1.00 (2.8%) | 1.00 (4.0%) | 2.00 (3.3%) |
| White | 16.0 (44.4%) | 10.0 (40.0%) | 26.0 (42.6%) |
| Other | 0 (0%) | 2.00 (8.0%) | 2.00 (3.3%) |
| Unknown | 1.00 (2.8%) | 0 (0%) | 1.00 (1.6%) |
| **Age** |  |  |  |
| Mean (SD) | 52.3 (16.9) | 55.3 (18.9) | 53.4 (17.5) |
| Median [Min, Max] | 52.0 [21.0, 89.0] | 59.0 [21.0, 84.0] | 54.0 [21.0, 89.0] |
| Missing | 7.00 (19.4%) | 8.00 (32.0%) | 15.0 (24.6%) |
| **AIS Grade at Discharge** |  |  |  |
| A | 5.00 (13.9%) | 2.00 (8.0%) | 7.00 (11.5%) |
| B | 2.00 (5.6%) | 1.00 (4.0%) | 3.00 (4.9%) |
| C | 4.00 (11.1%) | 5.00 (20.0%) | 9.00 (14.8%) |
| D | 22.0 (61.1%) | 14.0 (56.0%) | 36.0 (59.0%) |
| E | 1.00 (2.8%) | 1.00 (4.0%) | 2.00 (3.3%) |
| Unknown | 2.00 (5.6%) | 2.00 (8.0%) | 4.00 (6.6%) |
| **Neurological Level of Injury** |  |  |  |
| Cervical | 27.0 (75.0%) | 13.0 (52.0%) | 40.0 (65.6%) |
| Thoracic | 1.00 (2.8%) | 1.00 (4.0%) | 2.00 (3.3%) |
| Lumbar | 2.00 (5.6%) | 2.00 (8.0%) | 4.00 (6.6%) |
| Missing | 6.00 (16.7%) | 9.00 (36.0%) | 15.0 (24.6%) |
| **Injury Severity Score (ISS)** |  |  |  |
| Mean (SD) | 20.8 (8.61) | 23.3 (13.1) | 21.8 (10.6) |
| Median [Min, Max] | 17.0 [10.0, 45.0] | 21.0 [9.00, 75.0] | 19.0 [9.00, 75.0] |
| **Pain medications prior to SCI** |  |  |  |
| Yes | 5.00 (13.9%) | 6.00 (24.0%) | 11.0 (18.0%) |
| No | 31.0 (86.1%) | 19.0 (76.0%) | 50.0 (82.0%) |

**Figure 4.**
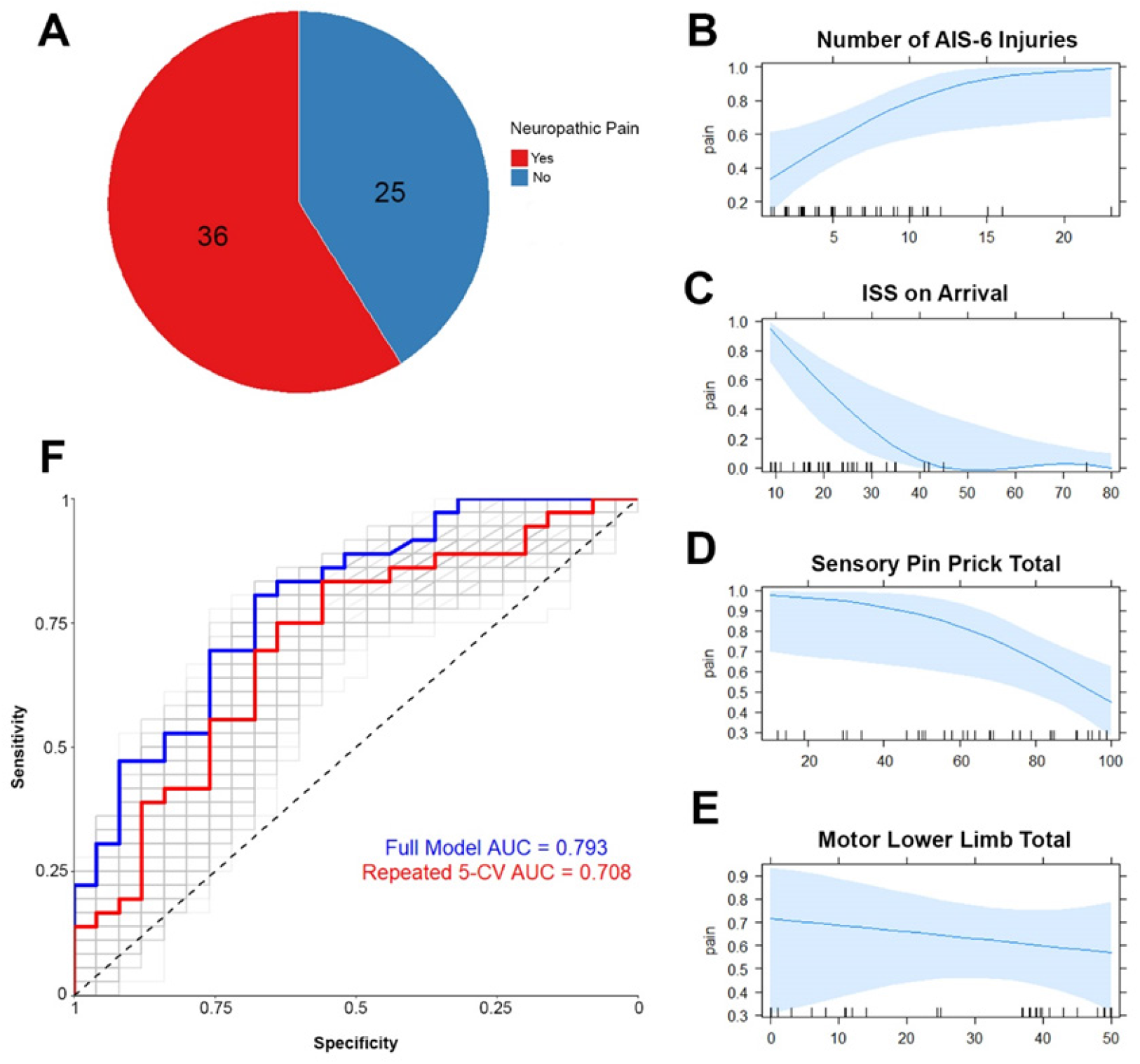
Logistic regression analysis suggests polytrauma and sensory scores as important predictors of neuropathic pain development after SCI. **A**. Out of 61 SCI patients that we have complete pain data, 36 reported they experience neuropathic pain at 6- and/or 12-months post SCI. The pain status (Yes/No) will be the dependent variable in the logistic regression model. **B-E**. Marginal effect plots of the four variables used in the logistic regression model. **F**. ROC analysis of the model. The full model including all 61 patients has an AUC of 0.793 (blue line). Repeated 1,000 times 5-fold cross validation has an average AUC of 0.708 (red line).

Although the drop is evident, the model still performs well, indicating that is not overfit on the data. Even so, an external validation of the model will be necessary to generalize our findings. In summary, our data and analysis suggest that commonly gathered variables at every level I trauma center at patient admission can be utilized to predict the development of neuropathic pain at the chronic stages after SCI.

## Discussion

Neuropathic pain is undeniably one of the most important ‘side-effects’ of SCI. There are several reports as well as anecdotes from SCI patients that prioritize the treatment of neuropathic pain over motor improvements in the long term (> 6 years).^11, 25-27^ Unfortunately, despite the great progress in the mechanistic understanding of neuropathic pain, reliable treatments are still elusive, and in most cases the patients will have to simply learn how to live and cope with the pain.^5^ One of best and recommended strategies for neuropathic pain management is early treatment.^28, 29^ Studies have shown that patients who are addressing their pain early show higher quality of life compared to those that begin treatments later.^30, 31^ To this end, our study sought to utilize a plethora of acute care hospital data from SCI patients and attempt to create a model predicting the development of neuropathic pain at 6- and 12-months post SCI.

Early prognostication of pain development will facilitate the earliest possible treatments for neuropathic pain and lead to significant higher life quality standards for SCI patients. Before initiating the acute care data analysis, we first looked at the TRACK-SCI enrolled patients database for the chronic pain status and how that is compared to previously reported summaries. From the 61 patients included in our analysis, 36 of them (59%) reported that they experienced neuropathic pain 6 and 12 months after SCI. From other similar studies, the neuropathic pain prevalence rate for SCI patients were 40%, ^32^ 57.4%, ^7^ 59%, ^8^ and 69.1%.^9^ Except for Kim et al,^9^ these figures are within the 38.58%-67.47% range proposed by Burke et al.^18^ That suggests that our cohort is well within the range of what is already known about neuropathic pain occurrence after SCI. While these studies agreed upon a similar neuropathic pain prevalence, no consensus was reached for which predictor(s) best suited to predict neuropathic pain. Marriage status, pain adaptation, cold-evoked dysesthesia or pain, old age, cause of trauma, type, and early onset of pain were suggested as possible biomarkers for prediction of chronic neuropathic pain.^7-9, 32^ While it is likely that the aforementioned variables could be predictive of pain development, our analysis revealed that acute polytrauma characteristics and motor and sensory scores from the ISNCSCI examination are additional top predictors of neuropathic pain development.

A caveat of our study is that the model was created and cross-validated on the same data from enrolled SCI patients of one hospital (ZSFG). While the 1,000 times repeated cross-validation strategy offers some confidence that the model is not overfitting on the data and hence leading to inaccurate conclusions, an independent validation cohort is of utmost importance. One of the greatest advantages of our proposed model is that it is composed of variables that are being collected routinely in every hospital setting. That could mean that as we continue to enroll more patients in TRACK-SCI that can act as an external validation cohort, colleagues at other centers could simultaneously quickly assess and validate or disprove our proposed model. Validating and expanding this model would have invaluable impact to the quality of life of SCI patients as it will enable clinicians to predict the development of chronic neuropathic pain during the first few days post SCI, thus opening a window of opportunity for preemptive treatments.

## Materials and Methods

### TRACK-SCI study data collection

All procedures for this study were conducted with the approval of the Human Subjects Review Boards at the University of California at San Francisco (UCSF) and the U.S. Department of Defense Human Research Protection Office. All English and non-English speaking patients who presented to the emergency department (ED) and were diagnosed with a traumatic SCI were initially eligible for the study. Patients who were < 18 years old, in-custody, prisoners, pregnant, or on medically indicated psychiatric hold were excluded. Informed consent was sought for all patients. For patients who were unable to sign for themselves due to their injury, a witness unaffiliated with the study was present throughout the consenting process and signed on the patient’s behalf. Patients incapable of consenting themselves were initially enrolled via a legally authorized representative (LAR; next of kin) or another suitable surrogate when one was available, then later approached for patient consent. Patients and surrogates had the option to participate in all or some of the following study portions: blood draws, ISNCSCI exams, and/or follow-up assessments. Patients were compensated ($50) after each time point (hospital stay, 3-month phone call, 6-month in-person visit, 12-month in-person visit) for a total of $200.

The foundation of the TRACK-SCI database is the NINDS-recommended common data elements (CDEs).^33^ Core CDEs are data elements that all SCI studies are strongly encouraged to use in collection of basic participant information. Additional measures from the International Spinal Cord Society (ISCoS) were also used. Data collection domains include demographic, clinical, radiologic, and functional outcome measures. All data collected from these CDEs were housed in a Research Electronic Data Capture (REDCap)^34^ database and include more than 21,000 data fields including additional institutional variables, calculated fields, repeated measures, date/time stamping of measures, and completion status log. Upon admission to the inpatient service, another 19,148 data fields regarding trauma characteristics, injury severity, blood pressure management, operating room procedures, interventions, hospital outcomes, high-frequency operating room vital signs, as well as motor-sensory exams and pain questionnaires are obtained from both paper and electronic medical records as well as participant interview.

REDCap is in full compliance with Health Insurance Portability and Accountability Act (HIPAA) security standards for protection of personal health information (PHI). The following CDE categories comprised the demographic and clinical data domain: (1) demographics, (2) health history, (3) injury-related events, (4) assessments and examinations. A total of 229 variables concerning patient demographics, medical history, and consent/contact information were collected through abstraction from electronic medical record systems and participant interviews.

The International Standard for Neurological Classification of Spinal Cord Injury (ISNCSCI)^35^ was used to assess motor and sensory function, and group patients by injury severity based on the ASIA impairment scale (AIS) which ranges from A (most severe – complete) to E (not impaired). ISNCSCI exams were conducted by trained personnel who completed the ASIA International Standards Training E Program (InSTEP) and in-person training. ISNCSCI exams were performed on all patients during the initial admission, either as part of clinical care if the treating provider completed InSTEP training, or separately for the research study if the ISNCSCI was not performed for clinical purposes. Occasionally, an ISNCSCI was not performed or not completed during the admission, usually because the patient was excessively sedated and could not participate in the exam. In the case of incomplete ISNCSCI exams, the assessor gave an estimated AIS grade based on the collected data and the overall clinical picture of the patient.

If possible, patients completed examinations at regular intervals including admission (day 0 = 0-23 hours from injury), every 24 hours until post-injury day 7, discharge, 6-month follow-up (+/-2 weeks), and 12-month follow-up (+/-2 weeks). All ISNCSCI exams results were included in the REDCap database.

### Data curation

All data curation and analysis was done in the R programming language^36^ using RStudio.^37^

#### Neuropathic pain data

For all chronic pain data we used the self-reported answers of TRACK-SCI enrolled patients on the International Spinal Cord Injury Pain Basic Data Set (ISCIPBDS) Version 2.0 and (2) the Douleur Neuropathique en 4 Questions (DN4) questionnaires. These questionnaires were administered over a phone interview at 3-, 6-, and 12-months post SCI. All patients were asked to describe their 3 worse pain problems and classify it as “neuropathic” or “nociceptive” after being provided appropriate definitions. According to the DN4 questionnaire a DN4 ≥ 4 classifies the pain as neuropathic. In most cases, all pain areas scored with DN4 ≥ 4 were indeed reported as neuropathic by the patients. In a handful of cases where the DN4 scores and the self-classification of the pain were not in agreement, self-classification was prioritized.

#### Acute care data

TRACK-SCI collects up to 22,000 data points per patient from hospital admission until discharge.^21, 38, 39^ All these data are stored in a RedCap database. For our analysis we excluded all high frequency vital monitoring variables (heart rate, mean arterial pressure, etc.) as well as all specific medications that patients were using before the SCI as well as the medications administered in the hospital. Pain medication prior to the injury was the only medication variable kept in our dataset. For functional outcomes we used the ISNCSCIs total subscores (motor and sensory) from the discharge examination. For the few patients who did not have an ISNCSCI examination at discharge we used their latest available at hospital.

### Non-linear principal component analysis

The final acute care dataset included 63 variables for 61 patients. The 61 patients were chosen based on the availability of pain data at 6- and/or 12-months after SCI. In order to perform principal component analysis to better understand how the variables are correlated with one another we had to deal with missing data. 4 o the 61 patients had missing data across the majority of the 63 variables and were removed from the subsequent steps. The remaining 57 patients had an overall 7.5% of missing data across all 63 variables. In order to proceed to the PCA we used multiple imputation to impute the missing values. We used the *mice*^*40*^ package in R to perform ten rounds of imputation with ten iterations each. The result was ten completed datasets with no missing values. Next, we merged these ten datasets by using the median imputed values for the numeric variables and the most frequent imputed level for the categorical variables. NL-PCA was performed using the *Gifi*^*41*^ package and the princals function in R. All visualizations and permutations tests were performed using the *syndRomics*^*42*^ package in R.

### Logistic regression

We used stepwise forward and backward logistic regression using the 20 variables from the 1,000 permutation tests of the communalities of the first 5 PCs with p < 0.1. The response variable was whether the patients reported neuropathic pain at 6- or 12-months post SCI. In order to classify a patient as experiencing neuropathic pain they had to self-report that one of their top 3 major pain problems was neuropathic. For patients that we had pain data for both 6- and 12-months post SCI we used the 12-month timepoint. The statistics and confusion matrix of the model were created using the *caret*^*43*^ package in R, the marginal effects plots with the *sjPlot*^*44*^ package, and the receiver operation characteristic analysis using the *ROCR*^*45*^ package.

## Data Availability

All data produced in the present study will become available upon publication of the manuscript in a peer-reviewed journal.

---

**Supplementary Figure 1.**
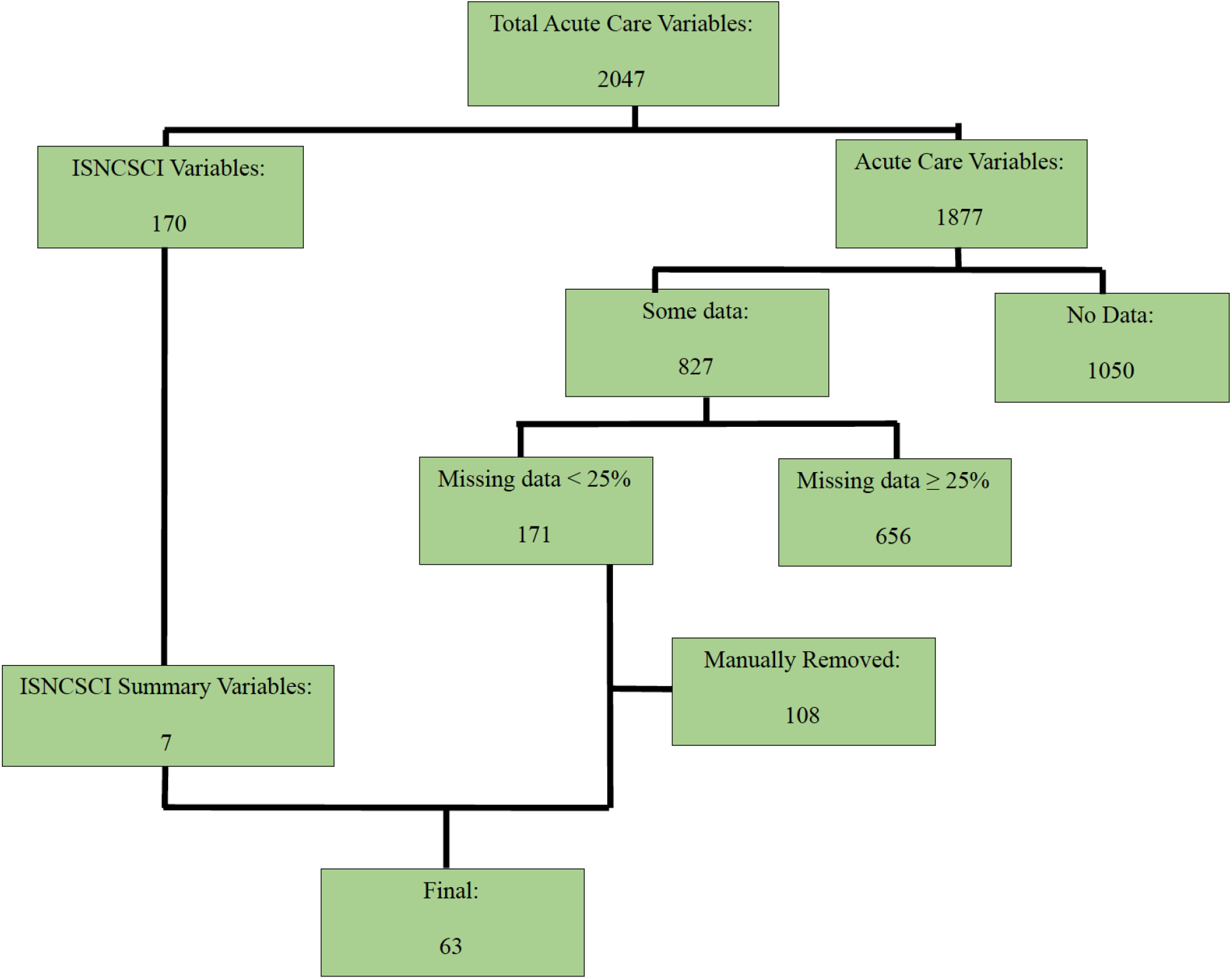
Flow chart of the acute care variable selection as potential predictors for the development of chronic neuropathic pain. Using the TRACK-SCI acute care database we started with 2,047 variables. By filtering out variables with missing data over 25%, manually removing variables the purpose of which was only for database maintenance, and by selecting only the total scores of the INSCSCI examination at discharge (or latest at hospital), the final number of variables used for multivariate analysis was 63.

**Supplementary Figure 2.**
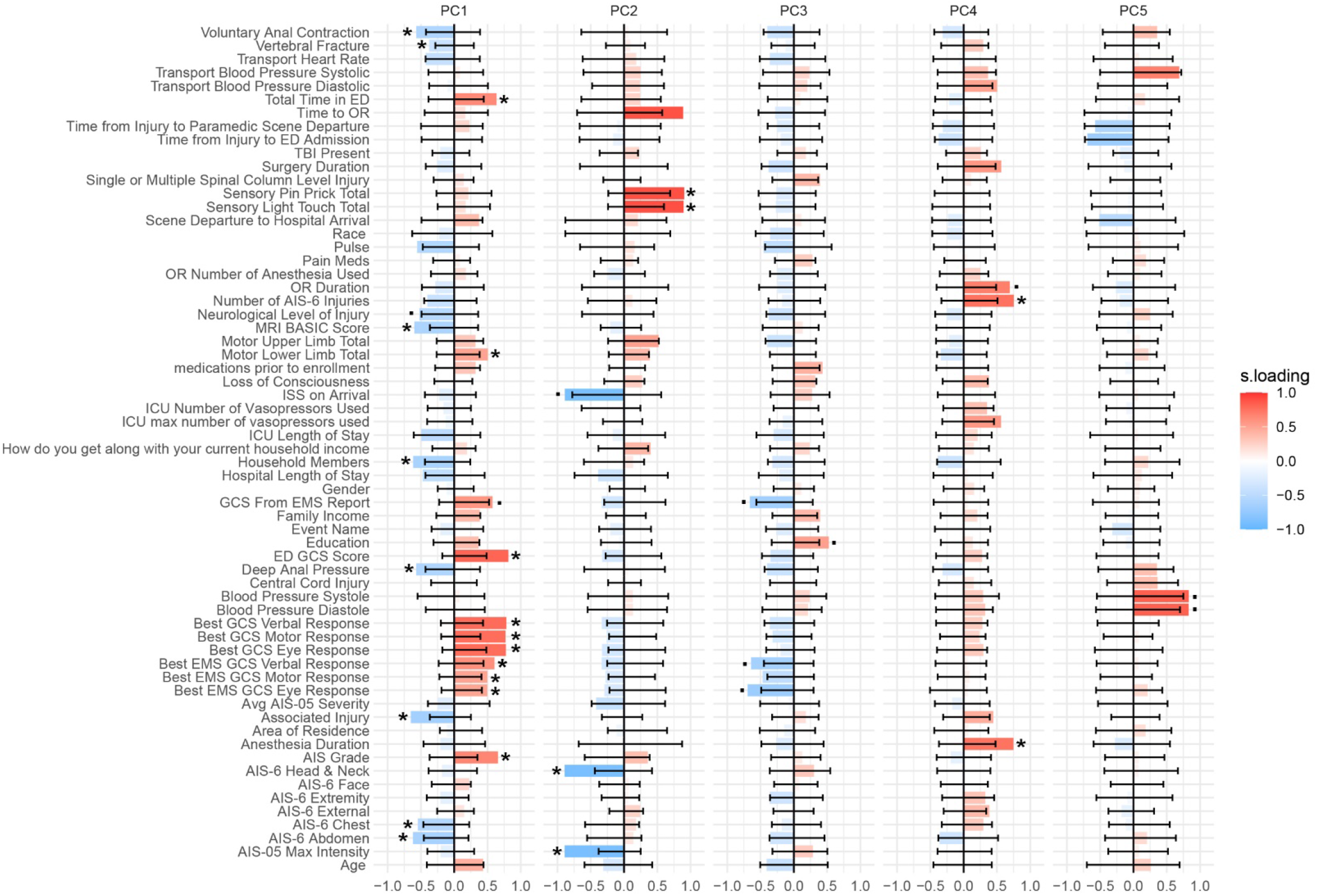
Barmap plot of the variable loadings of the first 5 PCs after 1,000 permutations for each variable x PC. * p < 0.05, p < 0.1.

**Supplementary Figure 3.**
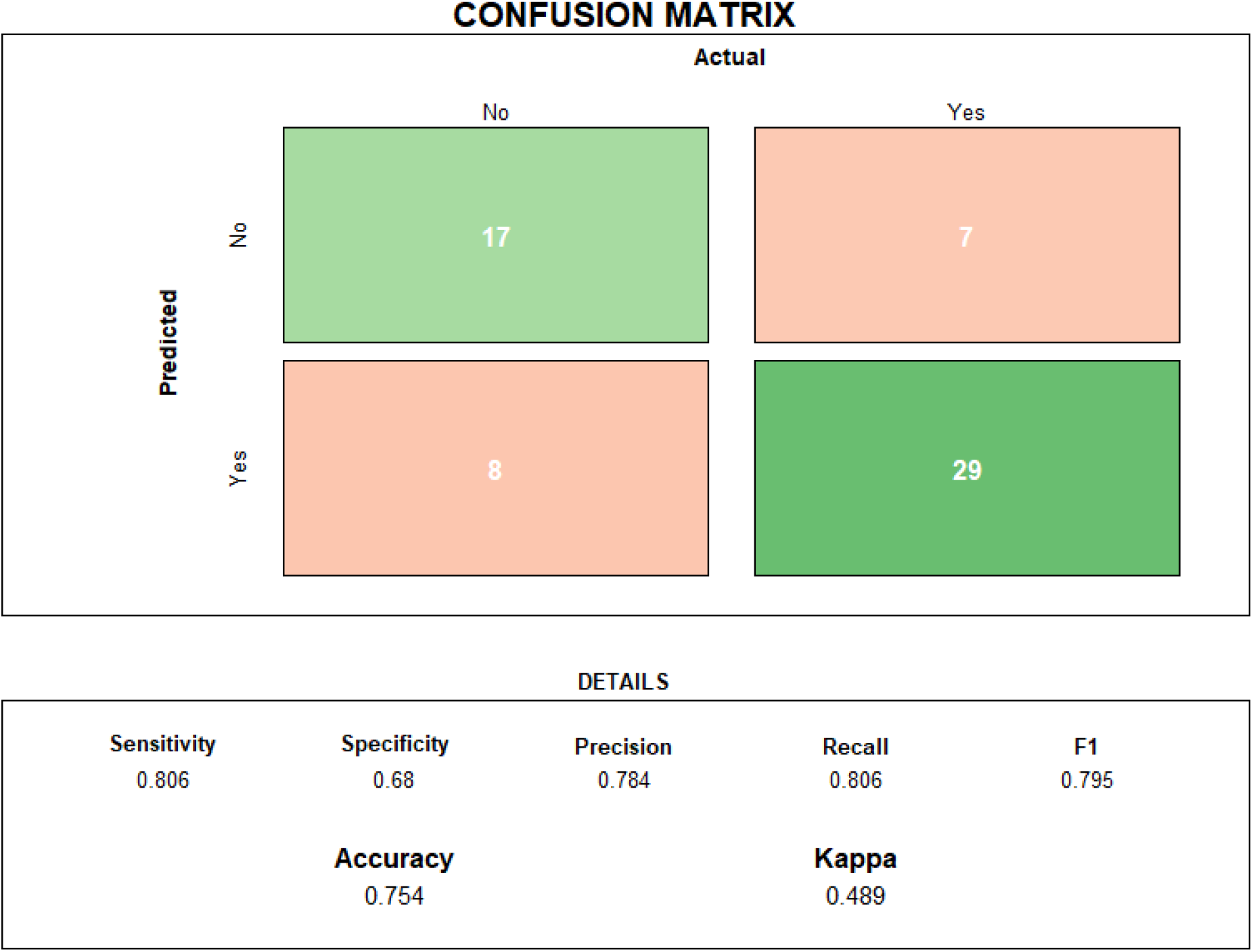
Confusion matrix and model metrics.

## Notes

### Competing Interest Statement

The authors have declared no competing interest.

### Funding Statement

The current work was supported by grants from the U.S. Department of Defense (W81XWH-13-1-0297 and W81XWH-16-1-0497) and Craig H. Neilsen Foundation (University of California, San Francisco, Spinal Cord Injury Center of Excellence special project award) to M.S. Beattie and an individual grant from Wings for Life to N. Kyritsis (WFL-US-07/18).

### Author Declarations

IRB of University of California San Francisco gave ethical approval for this work

